# Associations between screen use and antisocial behaviour in children and adolescents across development

**DOI:** 10.64898/2026.05.08.26352443

**Authors:** Natalia Tesli, Evgeniia Frei, Jaroslav Rokicki, Johan Siqveland, Alexey A Shadrin, Olav B Smeland, Ole A Andreassen

## Abstract

**Background:** Screen use is pervasive in childhood and adolescence, yet its role in antisocial behaviour (ASB) remains uncertain. While cross-sectional studies consistently link higher screen use to elevated ASB, longitudinal evidence is mixed, and few studies have controlled adequately for prior behaviour and genetic liability. Thus, it remains unclear whether these associations reflect prospective influences of screen exposure, or underlying vulnerabilities shared with ASB. We investigated whether screen use is a modifiable risk factor or a marker of underlying vulnerability.

**Methods:** We analysed data from up to 41,562 children in the Norwegian Mother, Father, and Child Cohort Study (MoBa). ASB traits and ICD-10-based conduct disorder (CD) diagnoses were assessed at ages 5, 8 and 14 years, together with screen use (total exposure and modality). Cross-sectional logistic regression models examined associations between screen use and ASB traits/CD at each age, adjusting for sex and parental education. Polygenic risk scores for ASB (PRS_ASB_) were used to assess genetic susceptibility and gene-environment interplay. Lagged logistic models tested whether screen use predicted later ASB, adjusting for prior ASB. Linear mixed-effects models examined developmental patterns across age.

**Results:** Higher screen use was positively associated with ASB traits and CD across all ages, with dose-response patterns across screen-use modalities. Social media showed the strongest modality-specific association at adolescence. In lagged models, screen use did not predict later ASB after adjustment for prior ASB. Longitudinal models showed significant but attenuating associations across development. PRS_ASB_ was independently and additively associated with ASB outcomes but did not interact with screen use.

**Conclusions:** We found that higher screen use was consistently associated with antisocial outcomes across childhood and adolescence. However, the absence of prospective associations after accounting for prior behaviour, together with independent genetic contributions, suggests that screen use may be better understood as a marker of underlying vulnerability rather than an independent driver of antisocial development.

## Introduction

Antisocial behaviour (ASB), encompassing aggression, rule violations, and delinquent acts, represents a major public health burden in child and adolescent mental health (McMahon and Frick, 2019). At the severe end of this spectrum lies conduct disorder (CD), a psychiatric diagnosis characterised by a persistent and impairing pattern of behaviour that violates societal norms and the rights of others (American Psychiatric Association, 2013). ASB and associated CD are relatively common in childhood and adolescence. Global prevalence of CD is estimated at up to 4% (Bachmann et al., 2024; Erskine et al., 2013), whereas subthreshold antisocial behaviours that do not meet diagnostic criteria are considerably more common (Reijneveld et al., 2012). Early manifestations of ASB are associated with an increased risk of persistent behavioural problems and a range of adverse adult outcomes, including impairments in social functioning, education attainment, and employment (Erskine et al., 2016; Rivenbark et al., 2018).

Given these long-term consequences, identifying risk factors for ASB is important for public health, with early intervention being critical for reducing the risk of persistent antisocial trajectories (Barker and Hawes, 2024; Farrington, 2005; Otto et al., 2021). Both genetic (Gard et al., 2019; Tielbeek et al., 2022) and environmental influences (Otto et al., 2021; Wesseldijk et al., 2018) contribute to the development and persistence of ASB traits. There is also growing interest in gene-environment interplay, whereby genetic liability may modify individual’s sensitivity to environmental exposures, potentially shaping antisocial outcomes.

Children’s use of digital devices is a new environmental exposure that has received increasing attention in recent years. Screen-based activities are now deeply embedded in daily life and account for a substantial proportion of children’s time from early childhood through adolescence (Mann et al., 2025; McArthur et al., 2022; Pew Research Center, 2024). Because screen use is both common and potentially modifiable, clarifying its relationship with ASB across development may have important implications for prevention.

A growing body of research has examined associations between children’s screen use and externalising behaviors (e.g., aggression, rule-breaking, and conduct problems), including outcomes across the antisocial spectrum, from subclinical ASB to diagnosis of CD. Many studies report that higher screen use is associated with elevated levels of externalising problems, particularly conduct-related symptoms, although findings vary by screen modality and study design. In a prospective cohort of 9–11-year-old children from the U.S.-based ABCD study, higher screen use was associated with an increased risk of new-onset parent-reported disruptive behavior disorders, with the strongest associations observed for social media use (Nagata et al., 2023), although the follow-up period was limited to one year. A longer longitudinal study following a sample of Canadian adolescents over five years identified associations between screen exposure and self-reported conduct problems, with effects varying by screen modality (Wallace et al., 2023). Similar patterns have also been reported in meta-analyses (Burkhardt and Lenhard, 2022; Eirich et al., 2022), including a recent meta-analysis of longitudinal studies of digital media use in children and adolescents (Teague et al., 2026). In particular, social media and video gaming were consistently associated with higher externalising problems, with video gaming additionally linked to aggression (Teague et al., 2026).

At the same time, emerging evidence suggests that associations between screen use and externalising behaviors may be bidirectional (Grund and Luciana, 2025; Neville et al., 2021). This raises the possibility that high screen use may also reflect underlying vulnerability to adverse mental health outcomes (Ayorech et al., 2023; Frei et al., 2025; Zhang et al., 2023), including ASB, rather than acting as a causal risk factor. Thus, the role of screen use and specific screen modalities in the development and persistence of ASB remains unclear, particularly with respect to temporal ordering and developmental changes. Addressing this question is particularly important given that ASB arises through a complex interplay of genetic liability and potentially modifiable environmental exposures.

Several limitations of the existing literature constrain inference. First, genetically informed designs examining the association between screen use and ASB remain scarce, meaning that associations reported in observational studies may partly reflect genetic confounding rather than environmental effects of screen exposure. Second, few studies have combined validated clinical diagnoses of CD with dimensional symptom measures of ASB. Third, even the longest longitudinal studies have been restricted to a single developmental period, typically preadolescence or adolescence, limiting insight into whether associations between screen use and ASB are stable across development or change in magnitude and nature across key developmental transitions.

The Norwegian Mother, Father, and Child Cohort Study (MoBa) (Brandlistuen et al., 2025) provides a unique opportunity to address these gaps. MoBa includes repeated assessments at ages 5, 8, and 14, and combines detailed information on children’s screen use with questionnaire-based measures of ASB, genetic data, and specialist healthcare diagnoses of CD. This design enables the examination of screen use across the full spectrum of antisocial outcomes, from dimensional symptoms to clinical disorder within a genetically informed longitudinal framework spanning key developmental stages.

The present study investigated cross-sectional and longitudinal associations between screen use and ASB across childhood and adolescence and evaluated the contribution of genetic liability and gene-environment interplay. Specifically, we examined whether screen use is more consistent with a modifiable risk factor for ASB or a marker of underlying vulnerability shaped by stable individual differences.

## Methods

### Study sample

The Norwegian Mother, Father and Child Cohort Study (MoBa) is a population-based pregnancy cohort study conducted by the Norwegian Institute of Public Health (Brandlistuen et al., 2025). Participants were recruited from all over Norway between 1999 and 2008. The women consented to participation in 41% of the pregnancies. The cohort includes approximately 114,500 children, 95,200 mothers and 75,200 fathers. The analytic sample for the present study included children with available questionnaire data at three assessment waves: ages 5, 8 and 14 years (maximum n = 41,562). MoBa is regulated under the Norwegian Health Registry Act. The current study was approved by The Regional Committees for Medical and Health Research Ethics (REK 2016/1226). Umbilical cord blood samples collected at birth were used for genotyping (Paltiel et al., 2014). Details of genetic data processing and quality control procedures have been described elsewhere (Corfield et al., 2024).

### Study variables

#### Conduct disorder (CD)

Data on CD diagnoses were obtained via linkage with the Norwegian Patient Registry (NPR), which includes diagnoses recorded in specialist healthcare (data coverage: 2008-2024). Nordic registries have demonstrated high validity for psychiatric diagnoses (Kouppis and Ekselius, 2020; Nesvåg et al., 2017). Individuals with at least one registered ICD-10 F91 diagnosis were classified as cases, regardless of comorbid psychiatric diagnoses. Controls were defined as individuals with no registered psychiatric diagnoses.

#### Antisocial behavioral (ASB) traits

ASB traits were assessed at ages 5, 8, and 14 years using DSM-based instruments capturing aggression, rule breaking, and conduct problems. At age 5, mothers answered selected questions from the Child Behavior Checklist for Ages 1½–5 (CBCL). At ages 8 and 14, ASB traits were measured with CD-related items from the Rating Scale for Disruptive Behavior Disorders (RS-DBD) based on maternal reports and self-reports, respectively (Supplementary Text 1, Table S1). Where relevant, ASB sum scores were dichotomised, defining “high ASB” as the top 10% at each age wave. For details see Supplementary Text 1.

#### Screen use behaviours

Screen use was assessed at ages 5, 8 and 14 years. At age 5, mothers reported the child’s average time spent watching TV and playing PC/TV games on typical weekdays and weekend days; a weighted weekly average was calculated. At age 8, mothers reported the child’s average weekday time spent watching TV/DVDs and playing video or computer games. At age 14, adolescents self-reported their average weekday time spent watching TV, playing games, and using social media. For details, see Supplementary Text 1 and Table S2.

#### Socioeconomic status

Parental highest level of education at baseline was used as a proxy for socioeconomic status. This variable was dichotomised into low (up to three years of high school) and high (college/university) level of education (see Supplementary Text 1).

#### Polygenic risk scores

Polygenic risk scores for ASB (PRS_ASB_) were derived from a genome-wide association study (GWAS) meta-analysis of broad ASB (Tielbeek et al., 2022) using PRSice2 (Choi and O’Reilly, 2019), applying a range of *p*-value thresholds (*P_T_* = 5 × 10^−8^, 10^−6^, 10^−5^, 10^−4^, 10^−3^, 0.01, 0.05, 0.1, 0.5, 1). To minimise multiple testing and capture genome-wide polygenic liability, a principal component summarising PRS across all *p*-value thresholds was extracted and used in the analyses (Coombes et al., 2020). Analyses were restricted to unrelated individuals of European ancestry to reduce bias due to population stratification.

### Statistical analyses

#### Cross-sectional models

At ages 5, 8, and 14 years, we used logistic regression to examine cross-sectional associations between screen use and antisocial outcomes. Screen use was operationalised as total exposure and standardised (z-scored) within each assessment wave to ensure comparability across ages. Two outcomes were examined, elevated ASB traits, defined as the top 10% within each wave, and CD diagnoses at the corresponding age. All models were adjusted for age, sex and parental education. Results are reported as odds ratios (ORs) with 95% confidence intervals (CIs). Modality-specific screen use (TV, gaming, social media) was also evaluated where available. Additionally, we estimated adjusted predicted probabilities of high ASB across screen use categories (see Supplementary Text 2).

To examine genetic liability, the models were re-estimated with PRS_ASB_ as a covariate, and a PRS_ASB_ × screen use interaction term was tested.

#### Sensitivity analyses

Sex-stratified models were fitted for cross-sectional associations with total and modality-specific screen use at each assessment wave, with high ASB traits as an outcome. Sex-stratified analyses were not conducted for CD due to low case counts.

#### Lagged models

To examine whether screen use was associated with subsequent antisocial outcomes, we fitted age-specific lagged logistic regression models for two intervals: ages 5-8 years and 8-14 years (see Supplementary Text 2). Antisocial outcomes at follow-up (dichotomised ASB traits or CD diagnosis) were regressed on screen use at the preceding wave, adjusting for baseline antisocial behaviour. Screen use and age were standardised within wave, and all models were adjusted for sex and parental education. Results are reported as ORs with 95% CIs. To examine the role of genetic liability, the lagged models were re-estimated with PRS_ASB_ as a covariate.

#### Longitudinal models

Associations between screen use and ASB traits across development were investigated using longitudinal linear mixed-effects models. Assessment wave (ages 5, 8, and 14) was treated as a categorical variable due to unequal spacing between intervals (3 and 6 years). Screen use was standardised within each wave and entered as both a main effect and in interaction with wave, allowing associations to vary across development. ASB sum scores were harmonised across waves by dividing sum scores by the number of items, yielding mean item scores. Models were adjusted for sex, age (centered within wave), parental education, and included a random intercept for repeated observations within individuals. This specification models within-individual associations across repeated measures while allowing associations to vary across developmental stages. Results are reported as regression coefficients (*β*) with 95% CIs. Alternative model specifications were explored but did not yield stable estimates; details are provided in Supplementary Text 2.

Subsequently, to investigate the role of genetic liability, PRS_ASB_ was included as a time-invariant covariate in longitudinal models. To assess potential bias due to selective attrition, models were re-estimated in a subsample with complete data across all assessment waves (n = 11,806). To account for multiple testing, false discovery rate (FDR) correction (Benjamini–Hochberg) was applied within predefined families of related cross-sectional hypotheses; lagged and longitudinal analyses were treated separately. Sensitivity analyses were considered exploratory and are reported with uncorrected *p*-values.

## Results

### Sample characteristics

Descriptive information on the main study variables and key demographic characteristics of the sample is presented in Table 1A. Screen use was common at all ages, with most children reporting approximately 1-2 hours daily use (Table 1B).

**Table 1.**
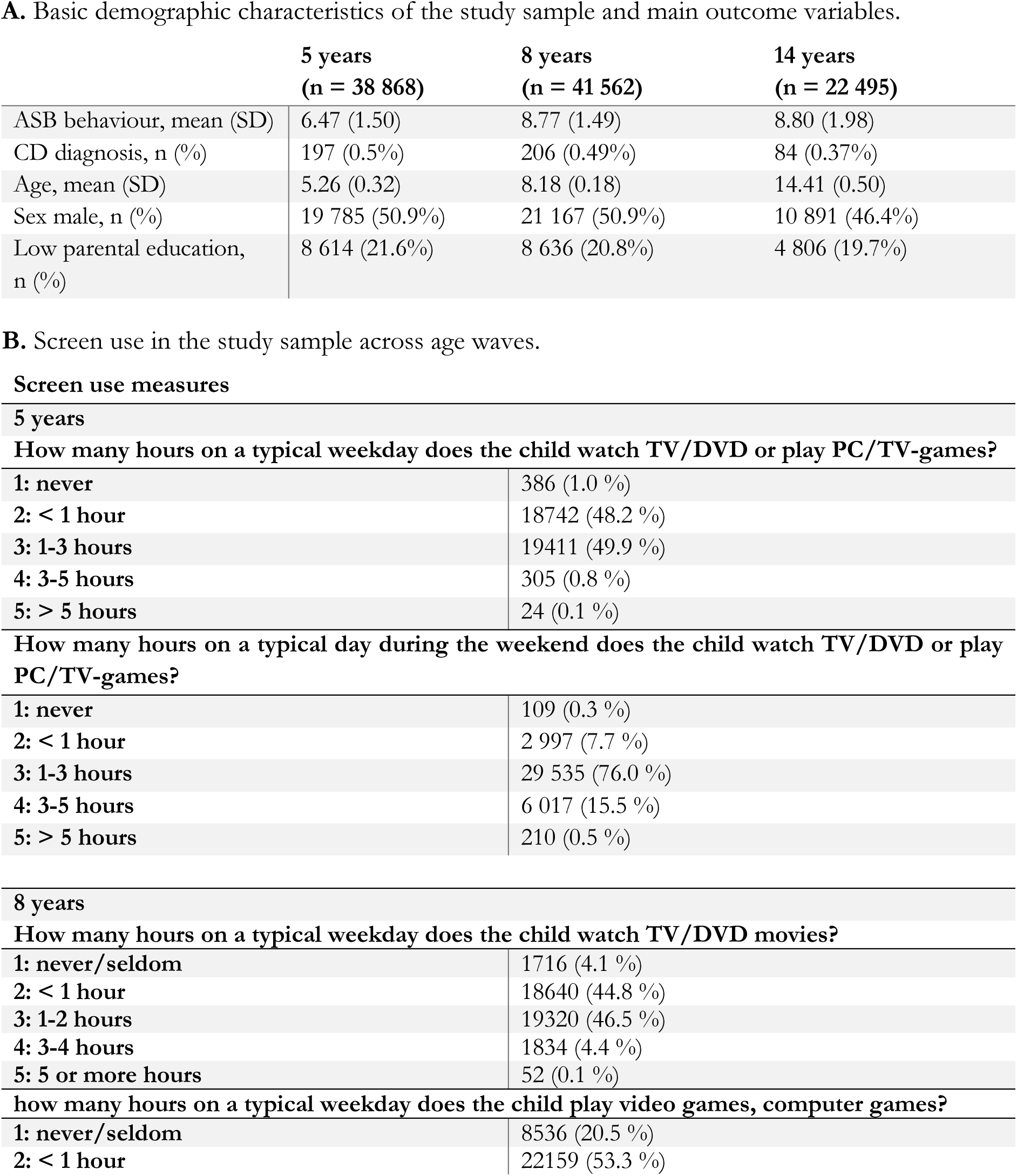
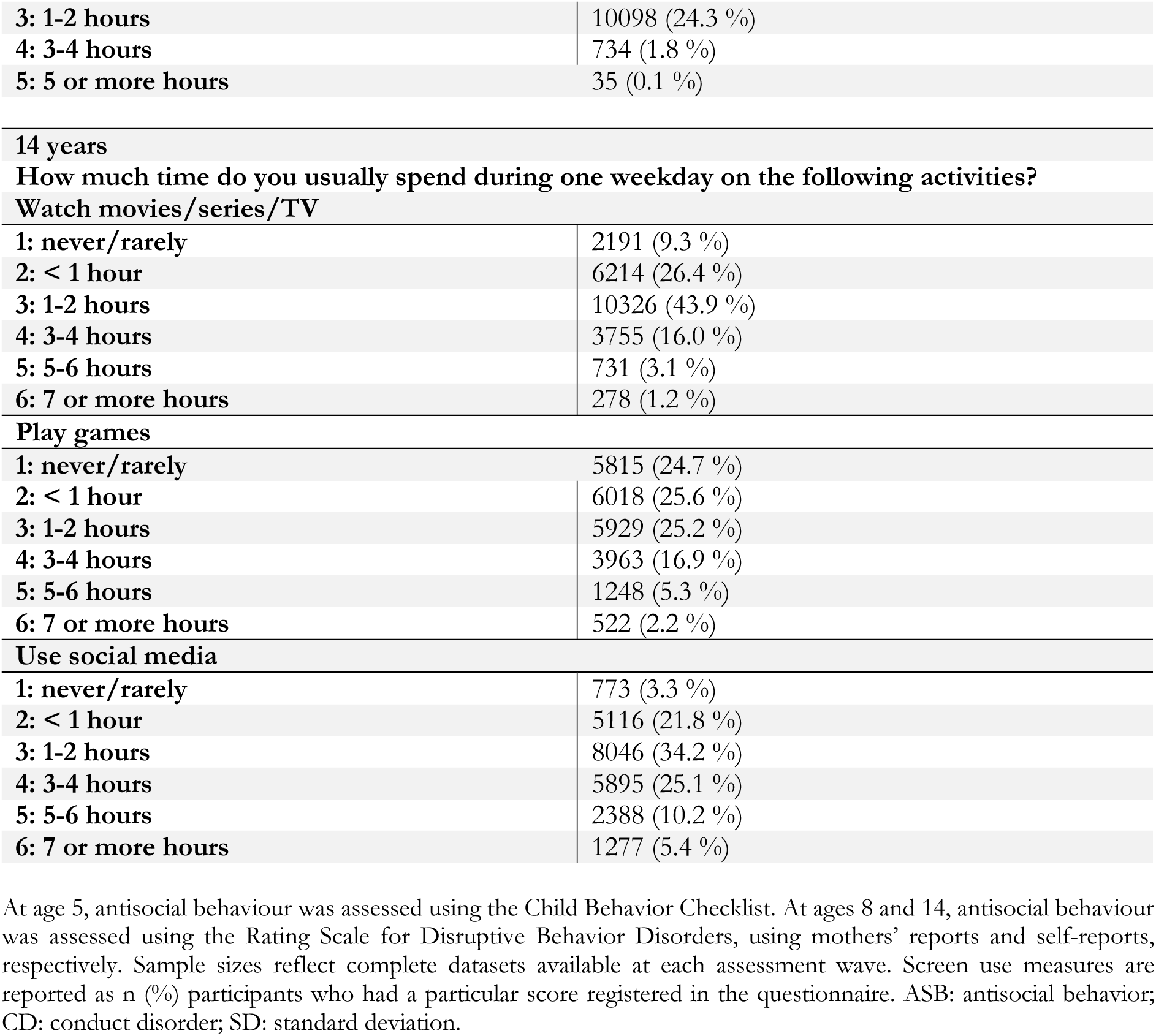
A. Basic demographic characteristics of the study sample and main outcome variables.

|  | **5 years**<br>**(n = 38 868)** | **8 years**<br>**(n = 41 562)** | **14 years**<br>**(n = 22 495)** |
| --- | --- | --- | --- |
| ASB behaviour, mean (SD) | 6.47 (1.50) | 8.77 (1.49) | 8.80 (1.98) |
| CD diagnosis, n (%) | 197 (0.5%) | 206 (0.49%) | 84 (0.37%) |
| Age, mean (SD) | 5.26 (0.32) | 8.18 (0.18) | 14.41 (0.50) |
| Sex male, n (%) | 19 785 (50.9%) | 21 167 (50.9%) | 10 891 (46.4%) |
| Low parental education, n (%) | 8 614 (21.6%) | 8 636 (20.8%) | 4 806 (19.7%) |

**How many hours on a typical weekday does the child watch TV/DVD or play PC/TV-games?**
|  |  |
| --- | --- |
| **1: never** | 386 (1.0 %) |
| **2: < 1 hour** | 18742 (48.2 %) |
| **3: 1-3 hours** | 19411 (49.9 %) |
| **4: 3-5 hours** | 305 (0.8 %) |
| **5: > 5 hours** | 24 (0.1 %) |

**How many hours on a typical day during the weekend does the child watch TV/DVD or play PC/TV-games?**
|  |  |
| --- | --- |
| **1: never** | 109 (0.3 %) |
| **2: < 1 hour** | 2 997 (7.7 %) |
| **3: 1-3 hours** | 29 535 (76.0 %) |
| **4: 3-5 hours** | 6 017 (15.5 %) |
| **5: > 5 hours** | 210 (0.5 %) |

**How many hours on a typical weekday does the child watch TV/DVD movies?**
|  |  |
| --- | --- |
| **1: never/seldom** | 1716 (4.1 %) |
| **2: < 1 hour** | 18640 (44.8 %) |
| **3: 1-2 hours** | 19320 (46.5 %) |
| **4: 3-4 hours** | 1834 (4.4 %) |
| **5: 5 or more hours** | 52 (0.1 %) |

**how many hours on a typical weekday does the child play video games, computer games?**
|  |  |
| --- | --- |
| **1: never/seldom** | 8536 (20.5 %) |
| **2: < 1 hour** | 22159 (53.3 %) |
| **3: 1-2 hours** | 10098 (24.3 %) |
| **4: 3-4 hours** | 734 (1.8 %) |
| **5: 5 or more hours** | 35 (0.1 %) |
| **14 years** |  |
| **How much time do you usually spend during one weekday on the following activities?** |  |
| **Watch movies/series/TV** |  |
| **1: never/rarely** | 2191 (9.3 %) |
| **2: < 1 hour** | 6214 (26.4 %) |
| **3: 1-2 hours** | 10326 (43.9 %) |
| **4: 3-4 hours** | 3755 (16.0 %) |
| **5: 5-6 hours** | 731 (3.1 %) |
| **6: 7 or more hours** | 278 (1.2 %) |
| **Play games** |  |
| **1: never/rarely** | 5815 (24.7 %) |
| **2: < 1 hour** | 6018 (25.6 %) |
| **3: 1-2 hours** | 5929 (25.2 %) |
| **4: 3-4 hours** | 3963 (16.9 %) |
| **5: 5-6 hours** | 1248 (5.3 %) |
| **6: 7 or more hours** | 522 (2.2 %) |
| **Use social media** |  |
| **1: never/rarely** | 773 (3.3 %) |
| **2: < 1 hour** | 5116 (21.8 %) |
| **3: 1-2 hours** | 8046 (34.2 %) |
| **4: 3-4 hours** | 5895 (25.1 %) |
| **5: 5-6 hours** | 2388 (10.2 %) |
| **6: 7 or more hours** | 1277 (5.4 %) |

At age 5, antisocial behaviour was assessed using the Child Behavior Checklist. At ages 8 and 14, antisocial behaviour was assessed using the Rating Scale for Disruptive Behavior Disorders, using mothers' reports and self-reports, respectively. Sample sizes reflect complete datasets available at each assessment wave. Screen use measures are reported as n (%) participants who had a particular score registered in the questionnaire. ASB: antisocial behavior; CD: conduct disorder; SD: standard deviation.

### Cross-sectional models

Total screen use was positively associated with ASB traits at all three assessment waves, with the strongest associations observed at 14 years (OR = 1.36, 95% CI 1.31-1.41). A similar pattern was observed for CD diagnosis (Figure 2A, Table S3), with somewhat larger effect sizes. The direction of the associations was consistent across waves and outcomes (ASB traits, CD diagnosis). The associations remained statistically significant after adjustment for sex, parental education, and age. The male sex and low parental education demonstrated consistently larger effects across models (Table S3).

**Figure 1.**
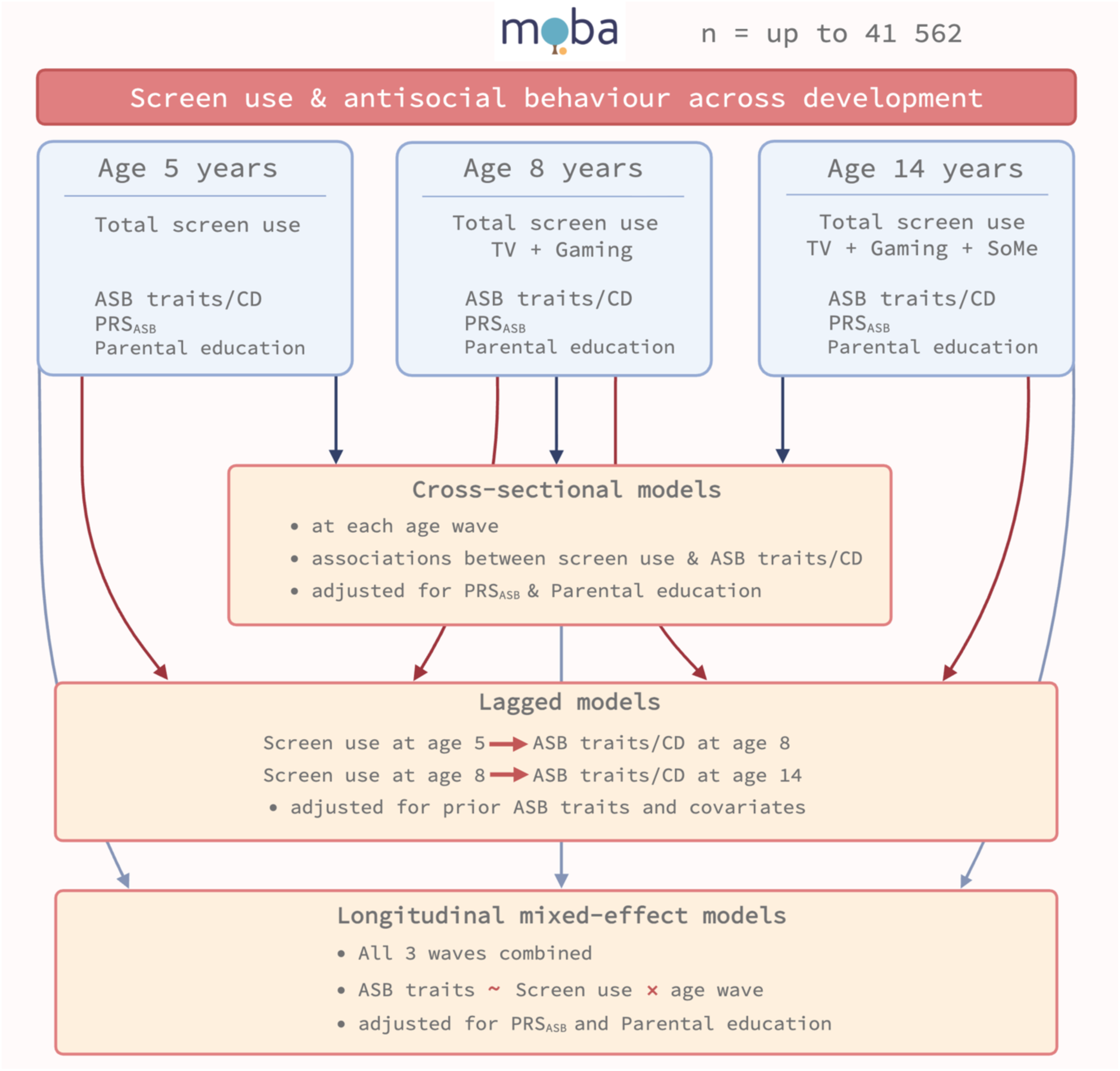
Study design and analytical framework. Data were drawn from up to 41,562 participants in the Norwegian Mother, Father and Child Cohort Study (MoBa) with repeated measures of screen use and antisocial behaviour at ages 5, 8, and 14 years. Screen use (TV, gaming, social media, where available by screen modality) and antisocial behaviour traits were assessed at each wave, together with covariates (sex, parental education) and polygenic risk scores for antisocial behaviour (PRSASB). Three complementary analytical approaches were applied: 1) cross-sectional models at each age, 2) lagged models examining temporal associations between screen use and later antisocial behaviour while adjusting for prior levels of antisocial behaviour, and 3) longitudinal mixed-effects models across all waves to evaluate developmental stability of associations.

**Figure 2.**
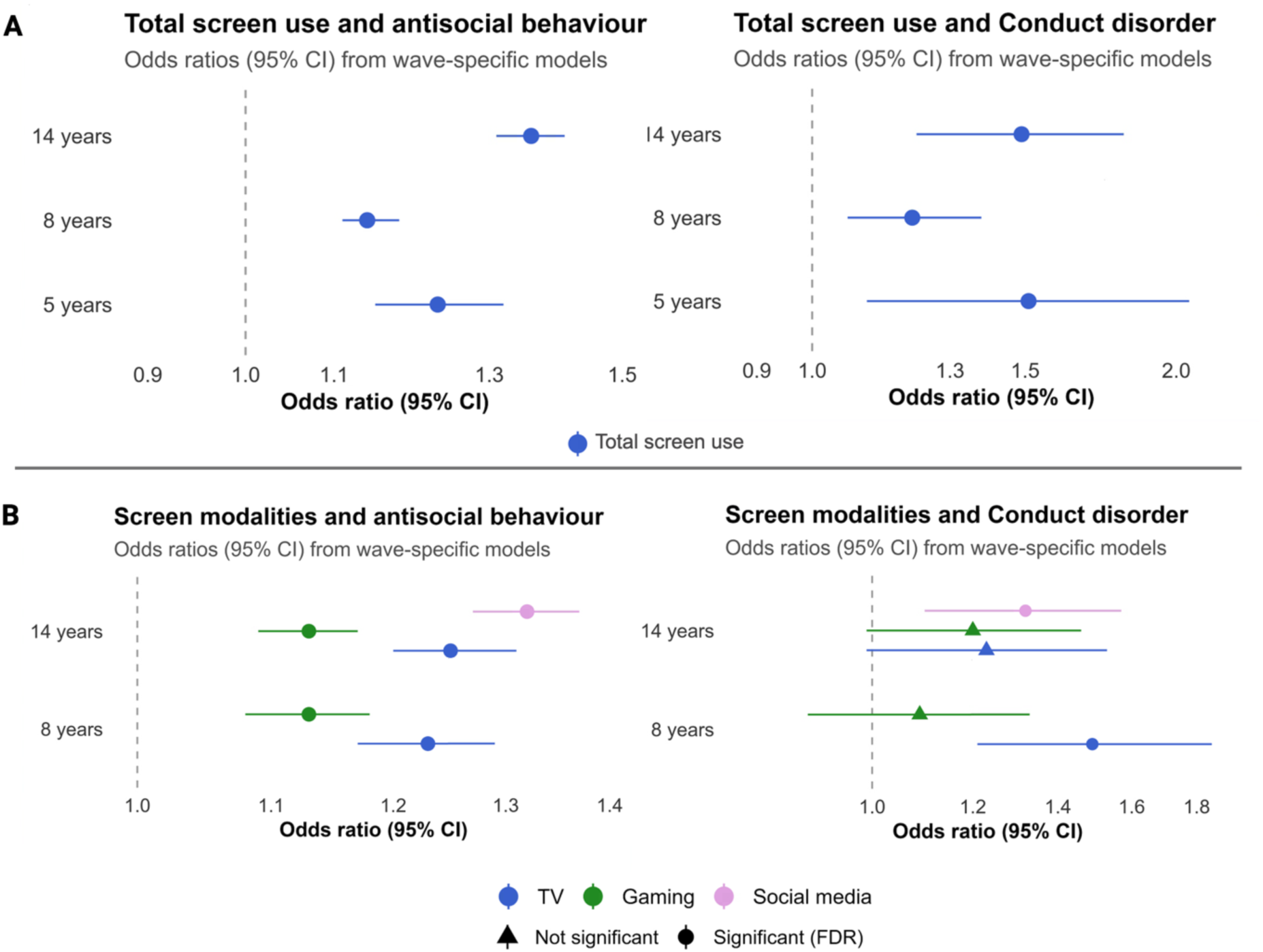
Cross-sectional associations between antisocial behavioural traits (left panel), conduct disorder diagnosis (right panel) and total screen use (A) and screen use modalities (B). The vertical line represents an effect estimate equal to zero. Error bars represent the 95% confidence intervals (CIs) of the estimate value. The logistic regression models were controlled for parental education, sex, and age. *P-values* were corrected with FDR.

In modality-specific analyses, TV viewing and gaming were positively associated with ASB traits at both ages, while social media use at age 14 showed the strongest association (Figure 2B). For CD diagnosis, effect estimates were generally in the same positive direction across modalities, but few remained statistically significant after correction for multiple testing, in contrast to the more consistent findings for ASB traits. Although effect sizes for CD were numerically larger than those observed for antisocial traits, CIs were wider, reflecting the smaller number of CD cases.

Adjusted predicted probabilities showed a consistent dose-response pattern across all assessment waves, with higher screen use categories associated with progressively higher probabilities of elevated ASB. The gradient was consistent across development and appeared more pronounced at age 14 (Figure 3).

**Figure 3.**
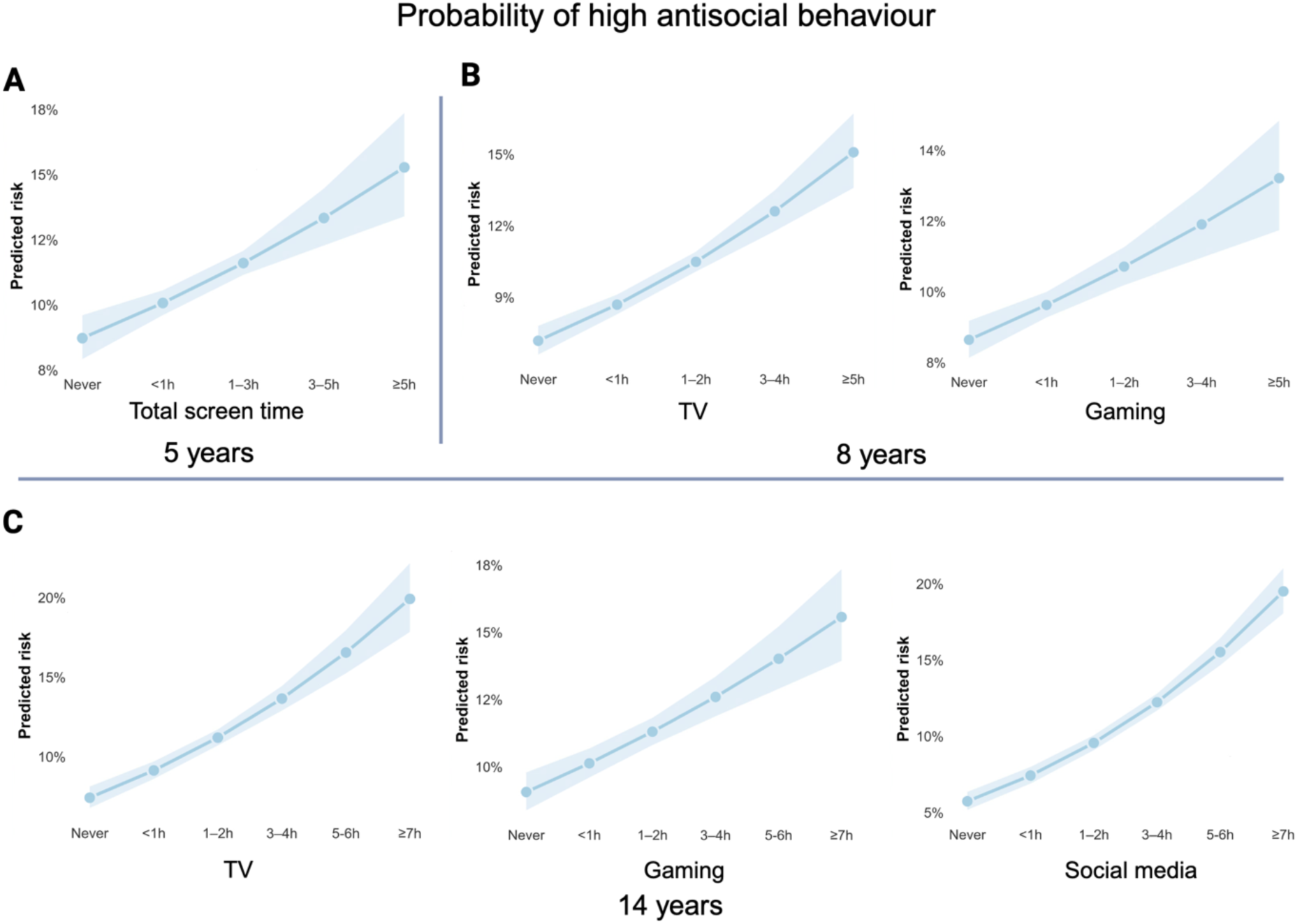
Adjusted predicted probabilities of high antisocial behaviour at age 5, 8 and 14 across screen use categories (in hours per day). Estimates were derived from logistic regression models adjusted for age, sex, and parental education, at ages 5 (A), 8 (B), and 14 years (C). High antisocial behaviour is defined as the top 10% within each wave. Shaded areas indicate 95% confidence intervals.

In models including PRS_ASB_, both screen use and PRS_ASB_ were independently associated with ASB traits across all age waves Inclusion of PRS_ASB_ did not attenuate the association between screen use and ASB traits (Figure 4). For CD diagnosis, PRS_ASB_ showed positive associations across waves. Adjustment for PRS_ASB_ resulted in modest attenuation of the screen use estimates for CD compared with models without PRS_ASB_, although effect estimates remained directionally similar. These findings suggest that genetic liability and screen use contribute largely additively to antisocial outcomes. Finally, no significant PRS_ASB_ × screen use interactions were observed at any age wave, indicating no evidence of multiplicative effect modification.

**Figure 4.**
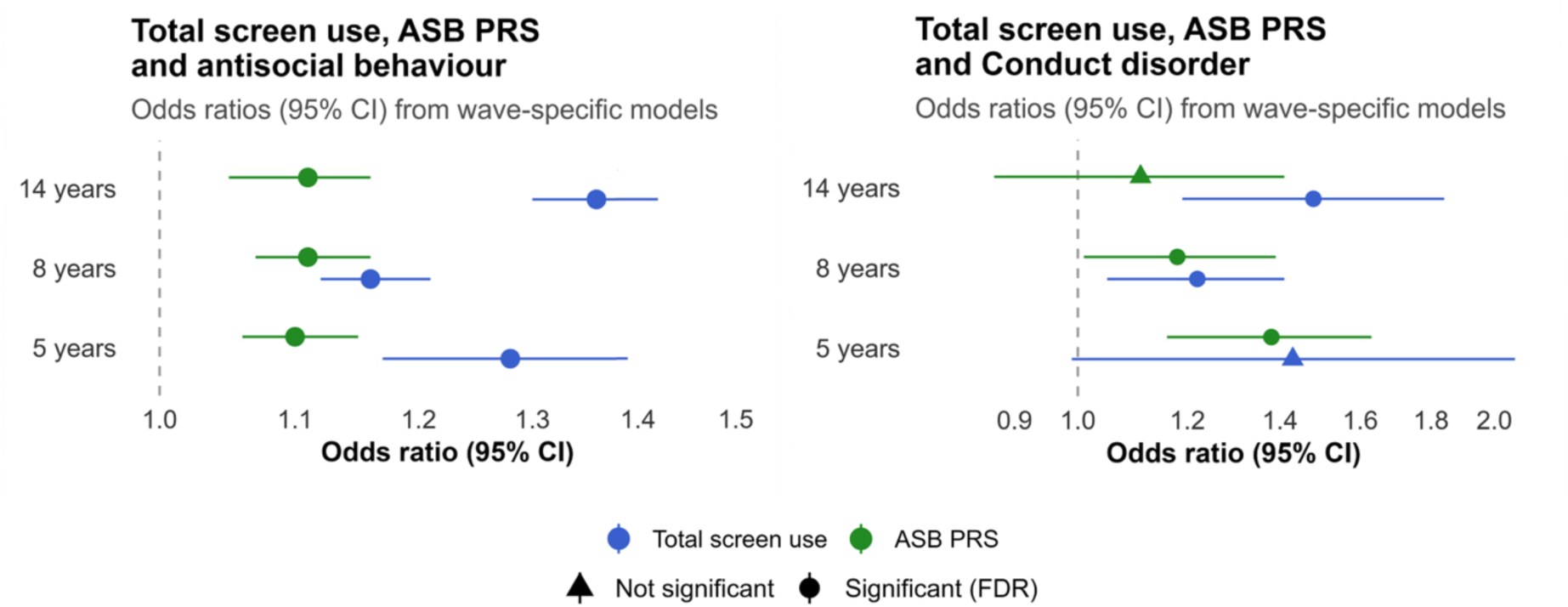
Cross-sectional associations between antisocial behavioural traits (left panel), conduct disorder diagnosis (right panel), screen use and polygenic risk score for antisocial behaviour at 5, 8 and 14 years. The vertical line represents an effect estimate equal to zero. Error bars represent the 95% confidence intervals (CIs) of the estimate value. The logistic regression models were controlled for parental education, sex, and age. *p-*values were corrected with FDR. ASB PRS polygenic risk score for antisocial behaviour.

### Sensitivity analyses

Sex-stratified analyses yielded patterns largely consistent with the main findings (Figure S1, Supplementary text 3). Positive associations between screen use and ASB traits were observed in both boys and girls across all age waves. There was no consistent sex difference in effect magnitude, although estimates were somewhat higher in girls at age 14, particularly for social media use (girls: OR = 1.42, 95% CI 1.34-1.52; boys: OR = 1.22, 95% CI 1.15-1.29). PRS_ASB_ remained independently associated with ASB traits in both sexes.

### Lagged models

Lagged models with dichotomised ASB traits provided no evidence that earlier screen use predicted subsequent ASB after adjustment for prior ASB and covariates (Table S4). From ages 5 to 8 years, screen use at age 5 was not associated with ASB at age 8 (OR = 1.03, 95% CI 0.99-1.07), whereas baseline ASB strongly predicted later ASB (OR = 5.62, 95% CI 5.11-6.19). A similar pattern was observed from ages 8 to 14 years, i.e., screen use at age 8 was not associated with ASB at age 14 (OR = 1.01, 95% CI 0.95-1.06) once prior ASB was accounted for. Across both developmental intervals, baseline ASB, male sex, and low parental education were significant predictors for later ASB.

Lagged models of CD diagnosis showed no evidence that earlier screen use predicted subsequent CD diagnosis after adjustment for prior CD diagnosis. Baseline CD was a very strong predictor of CD at follow-up, reflecting high temporal stability. Due to near-complete separation in these models, effect estimates for baseline CD were very large. The overall pattern indicated that screen us did not predict changes in CD beyond prior diagnosis.

### Longitudinal models

In mixed-effects models examining associations across three developmental waves, there was a significant positive association between total screen use and ASB traits, but this association varied across developmental waves (screen use × wave interaction, *p* < .001). Higher total screen use was associated with higher ASB traits at age 5 (*β* = 0.048, 95% CI 0.040 − 0.056, *p* <.001) (Figure S3). This association attenuated substantially across development. Interaction terms indicated that the screen-ASB association was weaker at ages 8 (*β*_interaction = -0.033, 95% CI -0.042 − -0.025, *p*<.001) and 14 years (*β*_interaction = -0.034, 95% CI -0.042 − -0.026, *p* <.001). Accordingly, the estimated association between screen use and ASB decreased to *β =* 0.015 at age 8 and to *β* = 0.014 at age 14.

In models including the PRS_ASB_, the magnitude and developmental pattern of the associations remained unchanged. PRS_ASB_ was independently associated with higher ASB across waves (*β* = 0.009, 95% CI 0.007 − 0.011, *p*<.001), indicating additive contributions of genetic liability and screen exposure.

Analyses restricted to children with complete data across all three waves (n = 11,806) yielded similar effect estimates and interaction patterns, demonstrating that the developmental attenuation of the screen use-ASB association was robust to selective attrition.

## Discussion

In this large population-based cohort spanning early childhood to adolescence, higher screen use was consistently associated with ASB across all ages, with clear dose-response patterns. However, these associations did not persist in prospective analyses once prior ASB was accounted for, and longitudinal models indicated attenuation of associations during development. PRS_ASB_ showed independent additive associations but did not interact with screen use. These findings suggest that screen use might be better understood as a marker of underlying liability rather than a causal risk factor for ASB.

The positive cross-sectional associations are consistent with prior work reporting modest but significant link between screen use and behavioural challenges (Eirich et al., 2022; Nagata et al., 2023; Neville et al., 2021; Teague et al., 2026; Wallace et al., 2023). In our study, the associations were stable across the full severity continuum of ASB, from self-reported traits to specialist-based CD diagnoses. Dose-response patterns support the robustness of these associations, also across development. Modality-specific analyses indicated that social media use showed the strongest associations with ASB traits at age 14, consistent with findings from other cohort studies (Nagata et al., 2023; Wallace et al., 2023). Although male sex was a strong overall predictor of ASB outcomes, sex-stratified analyses yielded broadly similar patterns in boys and girls, with somewhat stronger associations for social media use in adolescent girls.

However, screen use did not predict ASB outcomes; instead, prior ASB/CD strongly predicted later ASB. A recent meta-analysis found that many longitudinal studies did not adequately control for baseline behaviour while estimating screen use effects, and residual confounding of reported prospective associations cannot be ruled out (Teague et al., 2026). Indeed, effect sizes tend to be attenuated in studies that adjusted for prior behavioural differences (Eirich et al., 2022). Our findings are consistent with this pattern and with emerging longitudinal evidence that externalising behaviour predicts higher (Neville et al., 2021) or problematic (Grund & Luciana, 2025) screen use, but not vice versa. Additionally, prospective associations between screen use and mental health appear to vary by outcome domain, being somewhat weaker for externalising problems (Riehm et al., 2019). This supports the interpretation that associations between screen use and ASB may represent shared liability.

One might hypothesise that elevated screen use reflects underlying behavioural and temperamental characteristics, such as impulsivity, heightened reward sensitivity, or difficulties with self-regulation (Vasconcellos et al., 2025; Wallace et al., 2023), that could also contribute to ASB predisposition (Mann et al., 2018). Moreover, children with higher levels of behavioural dysregulation may be more likely to engage in screen-based activities, and their caregivers may be more likely to use screens as a strategy for managing stress or coping with challenging behaviour (McDaniel and Radesky, 2020; Radesky et al., 2023). In this context, high screen use can co-occur with ASB as a downstream correlate of shared liability rather than as a causal factor. Our findings also underscore the importance of both developmental stage, sex, and screen modality. For example, social media potentially provide a context in which relational aggression is more easily expressed among adolescents (Craig et al., 2020; Lin et al., 2025), which may partially explain its association with ASB and potential sex differences in these associations (Björkqvist, 2018; Lin et al., 2025). Nevertheless, these explanations remain hypothetical, and further research, especially more fine-grained and focused on specific subgroups or screen modalities, is needed to clarify which mechanisms account for the observed associations.

The results after inclusion of genetic data further supports a shared liability interpretation. PRS_ASB_ was associated with antisocial outcomes across all models, which indicates a robust contribution of genetic liability, consistent with previous work (Tesli et al., 2024). However, PRS_ASB_ did not substantially attenuate associations between screen use and ASB traits, and we found no evidence for PRS_ASB_ × screen use interaction. This pattern suggests mostly additive effects of genetic liability and screen exposure. Together with previous MoBa-based findings that psychiatric PRS are associated with patterns of screen use and that phenotypic associations partly reflect shared genetic influences (Frei et al., 2025), our results fit a model in which screen use and antisocial outcomes partly share underlying liability rather than a direct causal pathway.

From a developmental perspective, our findings further support this interpretation. Attenuation of associations between screen use and ASB across age appears to be inconsistent with an accumulating exposure model (Boxer et al., 2009; Wiedeman et al., 2015). Instead, it suggests that early individual differences, such as behavioural traits or family context, may shape both screen use and ASB, with their association diminishing as developmental contexts diversify. In addition, screen use becomes near-universal in adolescence (Smahel et al., 2020), reducing its discriminative value as an individual-level marker. Developmental changes in ASB, including the normative decline in overt conduct problems (Lansford, 2018; Moffitt, 1993), may further limit sensitivity to between-individual differences at older ages. Finally, the absence of consistent prospective associations aligns with population-level evidence indicating heterogenous trends in child mental health. Despite marked increases in screen use over the last decades, disruptive behavioural symptoms appear largely stable over time, while related domains such as impulsivity and hyperactivity show increases (Collet et al., 2026). Although such patterns cannot directly inform individual-level associations, they cautiously suggest that behavioural changes do not map straightforwardly onto rising screen use. This underscores the need for more fine-grained studies examining whether specific forms of screen use are differentially associated with distinct developmental outcomes, whether these associations are short-term or persistent, and whether they are more pronounced in vulnerable subgroups.

### Strengths and limitations

Some limitations should be acknowledged. First, screen use was assessed using self- and parent-reported measures, which are susceptible to bias (Parry et al., 2021), and did not capture content or context of screen activities. Second, while we adjusted for key confounders, residual confounding (e.g., broader family environment) cannot be ruled out. Third, selection bias and attrition may have influenced estimates (Biele et al., 2019), although results were consistent in complete-case sensitivity analyses. Finally, MoBa is conducted in a high-income Nordic society with strong social welfare systems, which may limit generalisability to other populations.

The study has several strengths. MoBa is one of the largest cohort studies to examine screen use and ASB. Repeated assessments span key developmental stages from early childhood to adolescence and capture important timepoints, including early exposure to digital media (McArthur et al., 2022), increasing salience of peer relationships (Rubin et al., 2011; Sørlie et al., 2021), and the emergence of clinically relevant mental health difficulties (Solmi et al., 2022). The combination of dimensional ASB traits and registry-based ICD-10 CD diagnoses enabled coverage across the full severity spectrum. In addition, the inclusion of PRSs provided a genetically informed perspective. Finally, the use of complementary analytic approaches enabled a nuanced evaluation of temporal ordering and developmental change.

### Conclusions

In the MoBa cohort, screen use was consistently associated with antisocial behaviour across development but showed no prospective effects and was attenuated over time. Together with independent genetic contributions, these findings suggest that screen use primarily reflects underlying liability rather than acting as a causal driver of antisocial development. While clinically informative as an indicator of risk, reducing screen use alone is unlikely to meaningfully alter antisocial trajectories without addressing broader behavioural and environmental factors.

## Acknowledgements

The Norwegian Mother, Father and Child Cohort Study is supported by the Norwegian Ministry of Health and Care Services and the Ministry of Education and Research. This research is part of the HARVEST collaboration, supported by the Research Council of Norway (#229624). The Norwegian Centre for Mental Disorders Research (NORMENT) provided genotype data, funded by the Research Council of Norway (#223273), Southeast Norway Health Authorities, and Stiftelsen Kristian Gerhard Jebsen. The Center for Diabetes Research, the University of Bergen, provided genotype data funded by the ERC AdG project SELECTionPREDISPOSED, Stiftelsen Kristian Gerhard Jebsen, the Trond Mohn Foundation, The Research Council of Norway, the Novo Nordisk Foundation, the University of Bergen, and The Western Norway Health Authorities.

This work was performed on Services for sensitive data (TSD), University of Oslo, Norway, with resources provided by UNINETT Sigma2 - the National Infrastructure for High Performance Computing and Data Storage in Norway.

## CrediT authorship contribution statement

**Natalia Tesli**: Conceptualisation, Data curation, Formal analysis, Visualisation, Writing – original draft; **Evgeniia Frei:** Data curation, Formal analysis, Writing – original draft, review & editing; **Jaroslav Rokicki:** Writing – review & editing; **Johan Siqveland:** Writing – review & editing; **Alexey A Shadrin**: Methodology, Writing – review & editing; **Olav B Smeland:** Writing – review & editing; **Ole A Andreassen:** Writing – review & editing, Resources, Funding acquisition.

## Data availability statement

Data from the Norwegian Mother, Father and Child Cohort Study is managed by the Norwegian Institute of Public Health. Access requires approval from the Regional Committees for Medical and Health Research Ethics (REC), compliance with GDPR, and data owner approval. Participant consent does not allow individual-level data storage in repositories or journals. Researchers seeking access for replication must apply via www.helsedata.no.

