## supplementary_materials for "Associations between screen use and antisocial behaviour in children and adolescents across development"

* Shared first authorship

Affiliations:

[a]Division of Mental Health Services, Akershus University Hospital, Lorenskog, Norway

[b]Centre for Precision Psychiatry, Division of Mental Health and Addiction, University of Oslo and Oslo University Hospital, Oslo, Norway

[c]Centre of Research and Education in Forensic Psychiatry, Oslo University Hospital, Oslo, Norway

[d]Department of Electronic Systems, Vilnius Gediminas Technical University, Vilnius, Lithuania

[e]Institute of Clinical Medicine, National Centre for Suicide Research and Prevention, University of Oslo, Oslo, Norway

**Supplementary Text 1.**

*Study Variables*

Specific questions used for ASB assessment are presented in Table S1. Internal consistency of the ASB scales was evaluated separately at each assessment wave using McDonald's omega based on polychoric correlations to account for the ordinal nature and skewed distributions of the items. Reliability was acceptable at age 5 ($\omega = 0.74$) and high at ages 8 and 14 ($\omega = 0.88$ and 0.89, respectively), indicating good internal consistency of the scale across development.

To facilitate interpretation and to allow for complementary modelling strategies, the ASB sum scores were additionally dichotomised to identify individuals with elevated ASB traits. A percentile-based threshold was applied, classifying children in the top 10% at each wave as having high ASB traits. This operationalisation enabled the use of logistic regression models in cross-sectional and lagged analyses, while preserving comparability across developmental stages.

Specific questions related to screen use are presented in Table S2. At age 5, a weighted weekly average was calculated as $5/7 \times$ weekday use $+ 2/7 \times$ weekend use. At age 8, the two items - average weekday time spent watching TV/DVDs and playing video/computer games were summed to derive a total screen use score. At age 14, the three items: average weekday time spent watching TV, playing games, and using social media were summed to derive a total screen use score.

Parental highest level of education, a proxy for socioeconomic status (SES), was extracted from the baseline MoBa questionnaire.  Education was measured on a six-point scale ranging from 1 (9-year elementary education) to 6 (college or university education > 4 years).

Detailed information about the MoBa questionnaires, including documentation, is provided on the Norwegian Institute of Public Health webpage: https://www.fhi.no/op/studier/moba/forskere/sporreskjemaer---mor-og-barn-unders/.

**Supplementary Text 2.**

*Statistical Analysis*

To facilitate interpretation of the logistic regression results, we estimated adjusted predicted probabilities of high ASB traits across categories of screen use at each assessment wave. These probabilities were derived from the fully adjusted logistic regression models (including sex, parental education and age as covariates) using marginal means (package *emmeans* in R) and were calculated on the response scale for each screen-use category while averaging over the observed distribution of covariates.

For longitudinal analysis, linear mixed-effects models with continuous antisocial behaviour scores were selected as generalised mixed-effects models with binomial outcomes did not yield stable estimates. Assessment wave (ages 5, 8, and 14) was treated as a categorical variable rather than as a continuous measure of age. This approach was adopted due to unequal spacing between intervals (3 and 6 years). Treating wave as categorical allowed more flexible estimation of associations at each developmental stage without imposing assumptions of linear change over time. Random slope models for screen use were also explored, however, due to limited within-individual variation in screen exposure, these models did not yield stable estimates. Final models included random intercepts only. Longitudinal analysis was performed with *lme4* package in R.

For lagged models, two model sets were estimated: (1) screen use at age 5 predicting antisocial outcomes at age 8 (n =27 061 complete cases), and (2) screen use at age 8 years predicting antisocial outcomes at age 14 years (n = 17 084 complete cases). In all lagged models, baseline antisocial behaviour was included as a covariate. Due to the low number of CD cases, models with CD diagnosis as the outcome were estimated using Firth's penalised logistic regression.

All statistical analyses were performed using R (version 4.2.3). To account for multiple testing while recognising the correlated structure of the outcomes, *p*-values were corrected within models examining total screen use in relation to antisocial behaviour traits and CD diagnosis, modality-specific screen use models, and models including genetic risk ($PRS_{ASB}$). Lagged and longitudinal mixed-effects models were treated as distinct developmental analyses addressing temporally ordered hypotheses.

**Supplementary Text 3.**

*Sensitivity analyses*

Sex-stratified analyses yielded patterns largely consistent with the primary models. At age 5, total screen use was positively associated with ASB traits in both boys and girls, with higher estimates observed in boys (boys: OR = 1.37, 95% CI 1.22-1.53; girls: OR = 1.16, 95% CI 1.02-1.33). At age 8, associations remained positive and comparable across sexes, and modality-specific analyses (TV and gaming) showed consistent effects in both boys and girls, with gaming showing higher estimates in girls (boys: OR 1.10, 95% CI 1.03-1.17; girls: OR = 1.29, 95% CI 1.16-1.42). At age 14, total screen use was associated with antisocial traits in both sexes. However, the effect estimates were larger in girls. Modality-specific models at age 14 showed positive associations for TV, gaming, and social media in both sexes, with the strongest estimates observed for social media in girls. Given lower baseline prevalence of antisocial traits among girls, apparent differences in magnitude between sexes should be interpreted cautiously. Across all age waves, $PRS_{ASB}$ remained independently associated with antisocial traits in both boys and girls.

### Supplementary Tables

**Table S1.** Specific questions used for the assessment of antisocial behavioural traits in the MoBa study.

| | |
|---|---|
| **Child Behaviour Checklist for Ages 1½–5**<br>**Age 5, maternal reports** | |
| To what extents are the following statements true of your child's behaviour during the last two months? | |
| Defiant | 1: Rarely/never<br>2: Sometimes<br>3: Often/typical |
| Doesn't seem to feel guilty after misbehaving | |
| Gets in many fights | |
| Hits others | |
| Punishment doesn't change his/her behaviour | |
| **Rating Scale for Disruptive Behaviour Disorders**<br>**Age 8, maternal reports** | |
| Mark the box that best describes your child's behaviour during the last 12 months/last year | |
| Bullies, threatens, or intimidates others | 1: Rarely/never<br>2: Sometimes<br>3: Often<br>4: Very often |
| Initiates physical fights | |
| Has been physically cruel to others | |
| Has harassed or injured animals physically | |
| Has stolen items of nontrivial value without confronting a victim (e.g. shoplifting) | |
| Has deliberately destroyed other's property | |
| Has been truant from school | |
| Has used an object that can cause serious physical harm to others (e.g. a bat, stone, knife, heavy toy) | |
| Mark the box that best describes your child's behaviour over the past 6 months | |
| Loses temper (tantrums) | 1: Rarely/never<br>2: Sometimes |
| Argues with adults | |
| Actively defies or refuses to comply with adults' requests or rules | |

| | |
|---|---|
| Deliberately annoys people | 3: Often<br>4: Very often |
| Blames others for his/her mistakes or misbehaviour | |
| Is touchy or easily annoyed by others | |
| Is angry or resentful | |
| Is spiteful or vindictive | |
| **Rating Scale for Disruptive Behaviour Disorders**<br>**Age 14, adolescent self-reports** | |
| Have you joined in or done any of this the past year? | |
| Bullied, threatened, or intimidated others | 1: Never/rarely<br>2: 1 time<br>3: 2-4 times<br>4: 5-10 times<br>5: 11-20 times<br>6: More than 20 times |
| Initiated physical fights | |
| Been physically cruel to others | |
| Harassed or injured animals physically | |
| Stolen items of nontrivial value without confronting a victim (e.g. shoplifting) | |
| Deliberately destroyed other's property | |
| Been truant from school | |
| Used an object that can cause serious physical harm to others (e.g. a bat, stone, knife, heavy toy) | |

**Table S2.** Specific questions used for the assessment of screen use in the MoBa study.

| | |
|---|---|
| **Age 5, maternal reports** | |
| How many hours does the child watch TV/DVD or play PC/TV-games? | |
| On a typical weekday | 1: Never<br>2: Less than 1 hour<br>3: From 1 up to 3 hours<br>4: From 3 up to 5 hours<br>5: 5 hours or more |
| On a typical day during the weekend | |
| **Age 8, maternal reports** | |
| How many hours on a typical weekday… | |
| … does the child watch TV/DVD movies? | 1: Never/seldom<br>2: Less than 1 hour<br>3: 1-2 hours<br>4: 3-4 hours<br>5: 5 hours or more |
| …does the child play video games, computer games, or handled video games? | |
| **Age 14, adolescent self-reports** | |
| How much time do you usually spend during one weekday on the following activities? | |
| Watch movies/series/TV | 1: Never/rarely<br>2: Less than 1 hour<br>3: 1-2 hours<br>4: 3-4 hours<br>5: 5-6 hours<br>6: 7 hours or more |
| Playing games (on PC, TV, tablet, mobile etc.) | |
| Communicating with friends on social media | |

**Table S3.** Logistic regression models for antisocial traits, conduct disorder diagnosis and screen use at 5, 8 and 14 years.

| | ASB traits | | | Conduct disorder | | |
|---|---|---|---|---|---|---|
| **At 5 years** | | | | | | |
| ***Predictor*** | ***OR*** | ***95% CI*** | ***p-value*** | ***OR*** | ***95% CI*** | ***p-value*** |
| **Screen time** | 1.23 | 1.15-1.32 | <0.001 | 1.51 | 1.11-2.05 | 0.008 |
| **Sex (male)** | 1.35 | 1.26-1.44 | <0.001 | 2.23 | 1.65-3.07 | <0.001 |
| **SES (low)** | 1.37 | 1.28-1.48 | <0.001 | 2.14 | 1.59-2.87 | <0.001 |
| **Age** | 0.97 | 0.94-1.00 | 0.69 | 1.12 | 0.96-1.29 | 0.15 |
| **At 8 years** | | | | | | |
| ***Predictor*** | ***OR*** | ***95% CI*** | ***p-value*** | ***OR*** | ***95% CI*** | ***p-value*** |
| **Screen time** | 1.14 | 1.11-1.18 | <0.001 | 1.21 | 1.07-1.38 | 0.004 |
| **Sex (male)** | 3.10 | 2.88-3.33 | <0.001 | 2.38 | 1.75-3.28 | <0.001 |
| **SES (low)** | 1.17 | 1.08-1.26 | <0.001 | 1.52 | 1.11-2.06 | 0.008 |
| **Age** | 1.00 | 0.91-1.04 | 0.8 | 1.06 | 0.92-1.36 | 0.5 |
| **At 14 years** | | | | | | |
| ***Predictor*** | ***OR*** | ***95% CI*** | ***p-value*** | ***OR*** | ***95% CI*** | ***p-value*** |
| **Screen time** | 1.36 | 1.31-1.41 | <0.001 | 1.49 | 1.22-1.81 | <0.001 |
| **Sex (male)** | 1.67 | 1.53-1.82 | <0.001 | 1.94 | 1.25-3.07 | 0.004 |
| **SES (low)** | 1.23 | 1.11-1.36 | <0.001 | 2.37 | 1.49-3.69 | <0.001 |
| **Age** | 1.07 | 1.02-1.11 | 0.002 | 1.07 | 0.86-1.32 | 0.5 |

The models included SES (parental education), sex, and age as covariates. *P-values* were corrected with FDR. OR: odds ratio, CI: confidence interval, ASB: antisocial behaviour.

**Table S4.** Lagged logistic regression models for binary antisocial behaviour outcomes (antisocial traits) covering two developmental intervals: 5 to 8 years and 8 to 14 years.

| Interval | Predictor | OR | 95% CIs | *p-value* |
|---|---|---|---|---|
| **5 → 8 years** | Screen use | 1.03 | 0.99 – 1.07 | 0.89 |
| **5 → 8 years** | Baseline ASB | 5.62 | 5.11 – 6.19 | <0.001 |
| **5 → 8 years** | Low parental education | 1.06 | 0.95 – 1.17 | 0.32 |
| **5 → 8 years** | Male sex | 3.21 | 2.93 – 3.52 | <0.001 |
| **8 → 14 years** | Screen use | 1.01 | 0.95 – 1.06 | 0.85 |
| **8 → 14 years** | Baseline ASB | 2.32 | 2.02 – 2.66 | <0.001 |
| **8 → 14 years** | Low parental education | 1.21 | 1.07 – 1.37 | 0.002 |
| **8 → 14 years** | Male sex | 1.42 | 1.28 – 1.57 | <0.001 |

OR: odds ratio, CI: confidence interval, ASB: antisocial behaviour.

### Supplementary Figures

**Figure S1.** Sex-stratified associations between antisocial behavioural traits and total screen use at 5, 8 and 14 years (upper panel); sex-stratified association between antisocial behavioural traits and modality-specific screen use at 8 and 14 years (bottom panel).

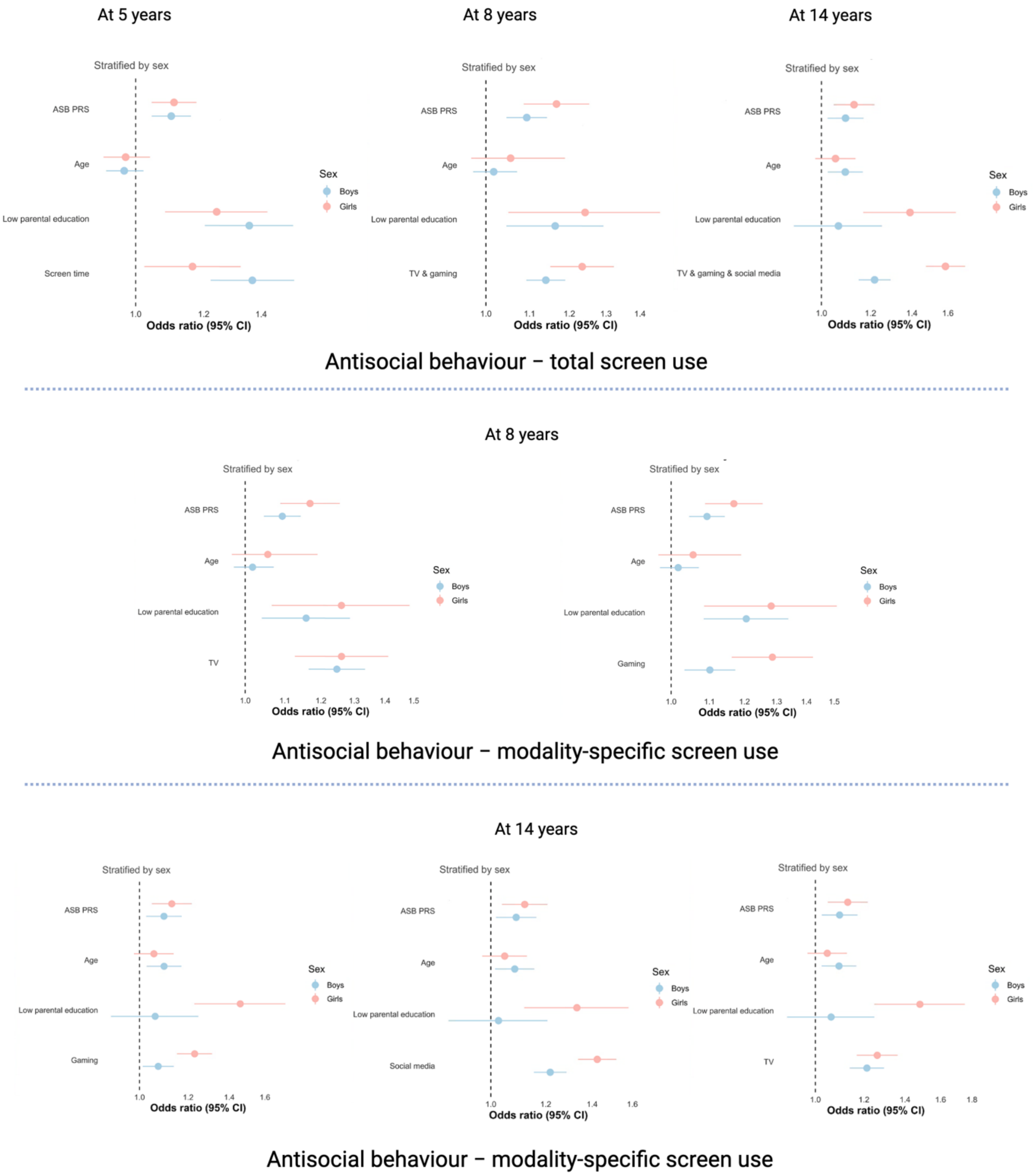

The vertical line represents an effect estimate equal to zero. Error bars represent the 95% confidence intervals (CIs) of the estimate value. The logistic regression models were controlled for parental education, sex, age and polygenic risk score for antisocial behaviour ($PRS_{ASB}$).

**Figure S2.** Predicted antisocial behaviour as a function of standardised screen use at ages 5, 8, and 14 years from longitudinal mixed-effects models.

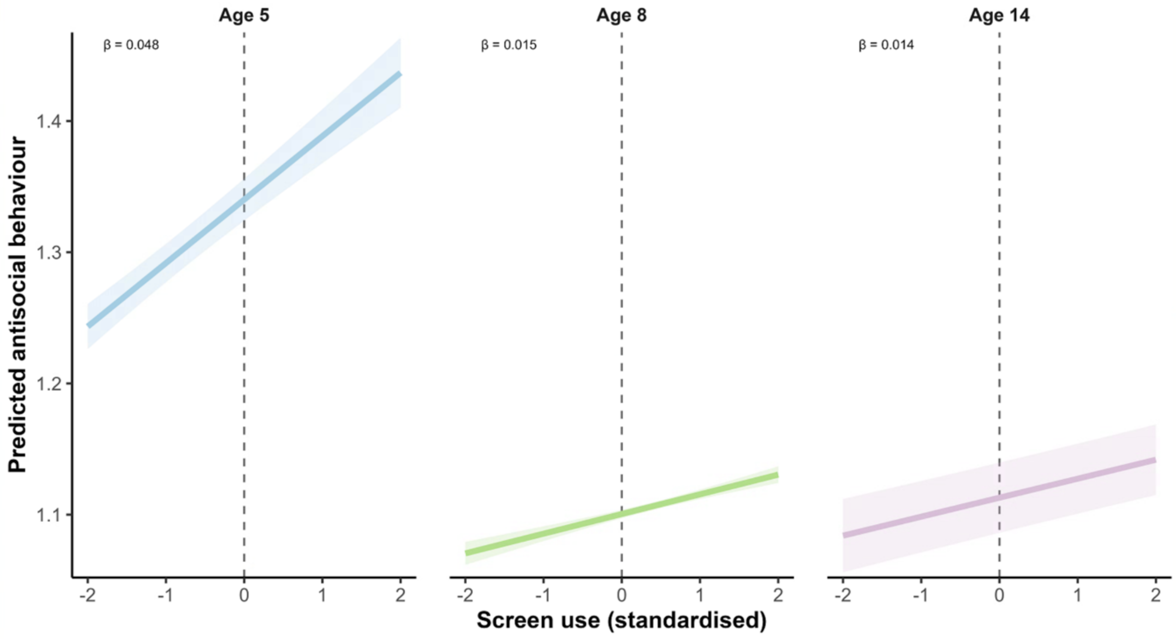

Solid lines show model-estimated associations within each wave, with shaded 95% confidence intervals. Wave-specific regression coefficients (*β*) are displayed in each panel.
